# Molecular Epidemiology, Risk Factors and Clinical Outcomes of Carbapenem- and Polymyxin-Resistant Gram-negative Bacterial Infections in Pregnant Women and Infants: A Systematic Review

**DOI:** 10.1101/2020.12.25.20248852

**Authors:** John Osei Sekyere, Melese Abate Reta

**Author notes:** Tweet: *“Clonal outbreaks of carbapenem- & colistin-resistant bacteria are common in neonatal intensive care units, causing very high mortalities. However, effective antibiotic therapy and infection control can prevent these deaths”.

## Abstract

**Background:** Carbapenems and polymyxins are last-resort antibiotics used to treat multidrug-resistant bacterial infections. However, resistance is increasing, even in vulnerable groups such as pregnant women and infants, for whom therapeutic options are limited.

**Method:** Using a diversity of databases, the literature was searched for studies investigating carbapenem and polymyxin resistance in pregnant women and infants (< 5 years).

**Result:** A final set of 73 manuscripts were used. In almost all countries, carbapenem/polymyxin-resistant *Klebsiella pneumoniae, Escherichia coli*, and *Acinetobacter baumannii* infect and/or colonizes neonates and pregnant women, causing periodic outbreaks with very high infant mortalities. Plasmid-borne *bla*_NDM_, *bla*_KPC_, *bla*_OXA-48_, *bla*_IMP,_ *bla*_VIM_ and *bla*_GES-5_ and ompK35/36 downregulation in clonal strains accelerate the horizontal and vertical transmission of carbapenem resistance in these pathogens. High prevalence of carbapenem/polymyxin resistance and carbapenemases were present in India, China, Pakistan, Thailand, Taiwan, Turkey, Egypt, Italy, USA, South Africa, Algeria, Ghana, and Madagascar. Factors such as antibiotic therapy, prolonged hospitalization, invasive procedures, mother/infant colonization, mechanical ventilation, low-birth weight and preterm state placed infants at high risk of carbapenem/polymyxin-resistant infections. Infant mortalities ranged from 0.2% to 36.8% in different countries.

**Conclusion:** Use of polymyxins to treat carbapenem-resistant infections is selecting for resistance to both agents, restricting therapeutic options for infected infants and pregnant women. However, appropriate infection control and antibiotic therapy can contain outbreaks and clear these infections. Antibiotic stewardship, periodic rectal and vaginal screening, and strict infection control practices in neonatal ICUs are necessary to forestall future outbreaks and deaths.

**Highlights:**

- Carbapenems & polymyxins are last-resort antibiotics used for multidrug-resistant infections
- Resistance to these two agents are reported in infants & pregnant women
- *K. pneumoniae, E. coli*, and *A. baumannii* are the most common pathogens
- Carbapenem & polymyxin resistance cause outbreaks with high infant mortalities
- Appropriate treatment & infection control can outbreaks & save lives

## Introduction

Resistance to antimicrobials remain a major challenge to public health, veterinary medicine, agriculture and aquaculture globally, owing to the limited or no therapeutic option available to treat multidrug-resistant (MDR) infections ^1–5^. To avoid depleting our antibiotics arsenals against infectious pathogens, certain agents are reserved as last-line drugs to fight difficult-to-treat infections ^6–8^. Currently, carbapenems, polymyxins (colistin) and tigecycline are used as reserved agents for MDR infections ^9,10^. However, the continual use of carbapenems and polymyxins to treat bacterial infections resistant to penicillins, cephalosporins, and carbapenems are increasing the rate of resistance to these last-resort antibiotics ^2,4,9,11^. The emergence of carbapenem- and/or polymyxin-resistant MDR infections is thus limiting therapeutic options and increasing hospital stay, healthcare costs, morbidity, and mortality ^12–14^. Subsequently, the WHO has classified carbapenem-resistant Gram-negative bacteria (CR-GNB) as critical priority pathogens for which novel antibiotics are urgently needed ^15,16^.

Pregnant women and infants (below 5 years old) are a vulnerable population with regards to bacterial infections due to their unbalanced and less matured immune system, respectively, and the limited therapeutic options available to them ^17–19^. Hence, the presence of MDR carbapenem and/or polymyxin-resistant infections among pregnant women and infants make them more susceptible to the ramifications of the infection ^13,19,20^. Whereas several studies have reported on pregnant women colonized with CR-GNB than infected by it, there is a risk of colonized pregnant women transferring these CR-GNB to their new borns ^19,21–24^. It is therefore not surprising that numerous studies report on CR-GNB- and polymyxin-resistant Gram-negative bacteria (PR-GNB)-infected neonates than colonized ones ^20,23,25,26^. Further, there have been abundant reports of clonal and polyclonal CR-GNB outbreaks in neonatal intensive care units (NICUs) globally, claiming the lives of several neonates as a result ^18,27–29^. This makes neonates more susceptible to CR-GNB and PR-GNB infections with their associated morbidity and mortality ^17–20,30^.

Notably, CR-GNB and PR-GNB carry other antibiotic resistance genes (ARGs) on mobile plasmids, transposons, integrons, integrative conjugative elements (ICE), and insertion sequences (ISs) that facilitate the horizontal transmission of these ARGs between species and clones ^13,20,31–33^. Specifically, extended-spectrum β-lactamases (ESBLs), fluoroquinolones, aminoglycosides, tetracycline, sulphamethoxazole-trimethoprim, and macrolide ARGs are also found in CR-GNB and PR-GNB. Hence, under antibiotic selection pressure, MDR plasmids and clones proliferate, leading to outbreaks ^10,27,34,35^. It is thus not surprising that most neonatal CRE infections occur in NICUs during clonal outbreaks in which the isolates harbour multiple ARGs on mobile plasmids ^20,25,26,30,36,37^. Analyses of the data obtained from this study confirms this observation.

### Evidence before this review

A thorough review of the literature showed the absence of a review addressing carbapenem and polymyxin resistance among pregnant women and neonates. Only one review addressed carbapenem-resistance among Gram-negative bacteria (GNB) in children^14^ whilst another only addressed CRE-causing neonatal sepsis in China^38^. Thus, this is the first work to systematically review and statistically analyse the literature on carbapenem and polymyxin resistance among pregnant women and infants.

### Literature & databases search strategy

All literature published in English were searched on Medline/PubMed, Web of Science, ScienceDirect, HINARI, Cochrane library electronic databases, and Google scholar using the following search words: “Gram-negative bacteria”, “epidemiology”, “prevalence”, “incidence”, “risk factors”, “determinant”, “associated factors”, “carbapenem”, “carbapenem resistance”, “colistin”, “polymyxin resistance”, “carbapenem-resistant Gram-negative bacteria”, “colistin-resistant Gram-negative bacteria”, “carbapenem resistance determinants”, “carbapenem resistance genotypes”, “carbapenem resistance mechanisms”, “colistin resistance determinants”, “colistin resistance genotypes”, “colistin resistance mechanisms”, “infants’’, “neonates”, “children” and “pregnant women”. These search words were further paired with each other in a factorial fashion and search strings were implemented using “AND” and “OR” Boolean operators. “OR” was used between “colistin” and “polymyxin” as in “colistin OR polymyxin”. No other filters were employed and the search was carried out up to 30^th^ September 2020. Both authors undertook the literature search, title and abstract screening and resolved outstanding discrepancies by consensus, using the inclusion and exclusion criteria.

### Inclusion and exclusion criteria

All articles addressing the following were included: 1) molecular testing of carbapenem- and polymyxin (colistin)-resistant GNB isolated from pregnant women and/or infants; 2) phenotypic antimicrobial sensitivity testing (AST) of carbapenem and polymyxin (colistin)-resistant GNB isolated from pregnant women and/or infants; 3) epidemiology (incidence, prevalence, risk factors and/or clinical outcomes) of carbapenem and/or polymyxin (colistin) resistance in pregnant women and/or infants (<5years old); 4) articles on pregnant women and infants with carbapenem or polymyxin (colistin)-resistant infections or colonization. Articles reporting on carbapenem-and/or polymyxin (colistin)-resistant GNB isolated from non-pregnant women and/or infants greater than 5 years old were excluded.

### Statistical analyses

Data on isolation country, year of isolation, carbapenem and polymyxin resistance profiles, GNB species and clones, carbapenem and polymyxin resistance mechanisms, age, sex, clinical history, colonization or infection, clinical history, previous and current medications, clinical outcomes, mortality rate and risk factors were extracted from the included articles and populated into Microsoft Excel. Further analyses were undertaken to compute isolation rates, prevalence, resistance rates, and mortality rates per country (Tables S1-S3).

## Results

### Included studies and samples

A total of 1427 articles were retrieved from the various databases, of which 73 studies from 28 coutries worldwide were finally included for the qualitative and quantitative analyses (Figure S1). Studies describing carbapenem resistance in infants (n = 65 studies) were from 24 countries, involved 49 154 infants (< 5 years), and resulted in the isolation of 11 441 GNB cultures with 23.3% isolation rate (Table S1). Carbapenem resistance in pregnant women (n = 7 studies) were reported from six countries among 1892 pregnant women; this resulted in 285 GNB cultures and an isolation rate of 15.1% (Table S2). Included studies on polymyxin resistance (n = 7) were mainly in infants (n = 16 556), with one being from a pregnant woman ^39^; 1345 GNB cultures were obtained, yielding an isolation rate of 8.1% (Table S3) (Figure S2).

Fifty studies on infants involved carbapenem resistance infections, 11 involved colonization by CR-GNB, and four involved both infection and colonization. At least 22 studies out of the 50 describing GNB infections in neonates involved sepsis cases (Table S1). Notably, all the studies on PR-GNB in infants also involved infection cases with no colonizations (Table S3). However, six of the seven studies on CR-GNB in pregnant women were mainly colonization cases with no infections, with only one study being an infection case in the USA (Table S2). There were higher GNB isolations from India, Bangladesh, Pakistan, Morocco, China, Turkey, South Africa, USA, and Cambodia than from the other countries (Figure S2).

### Species, clones, and resistance rates

On a per country basis, *Escherichia coli, Klebsiella pneumoniae, Acinetobacter baumannii, Enterobacter sp*. (cloacae), and *Pseudomonas aeruginosa* were the most dominant species causing infections or colonization in infants and pregnant women, with *Citrobacter sp*., *Proteus sp*., *Morganella sp*., *Serratia sp*., and *Salmonella sp*. being some of the less common species (Figure 1). Although these species differed in terms of prevalence per country, *E. coli, K. pneumoniae* and *A. baumannii* were the most common in almost all countries whilst *Pseudomonas aeruginosa* were highly prevalent in Italy, Nepal, USA, Turkey, and Kenya.

**Figure 1.**
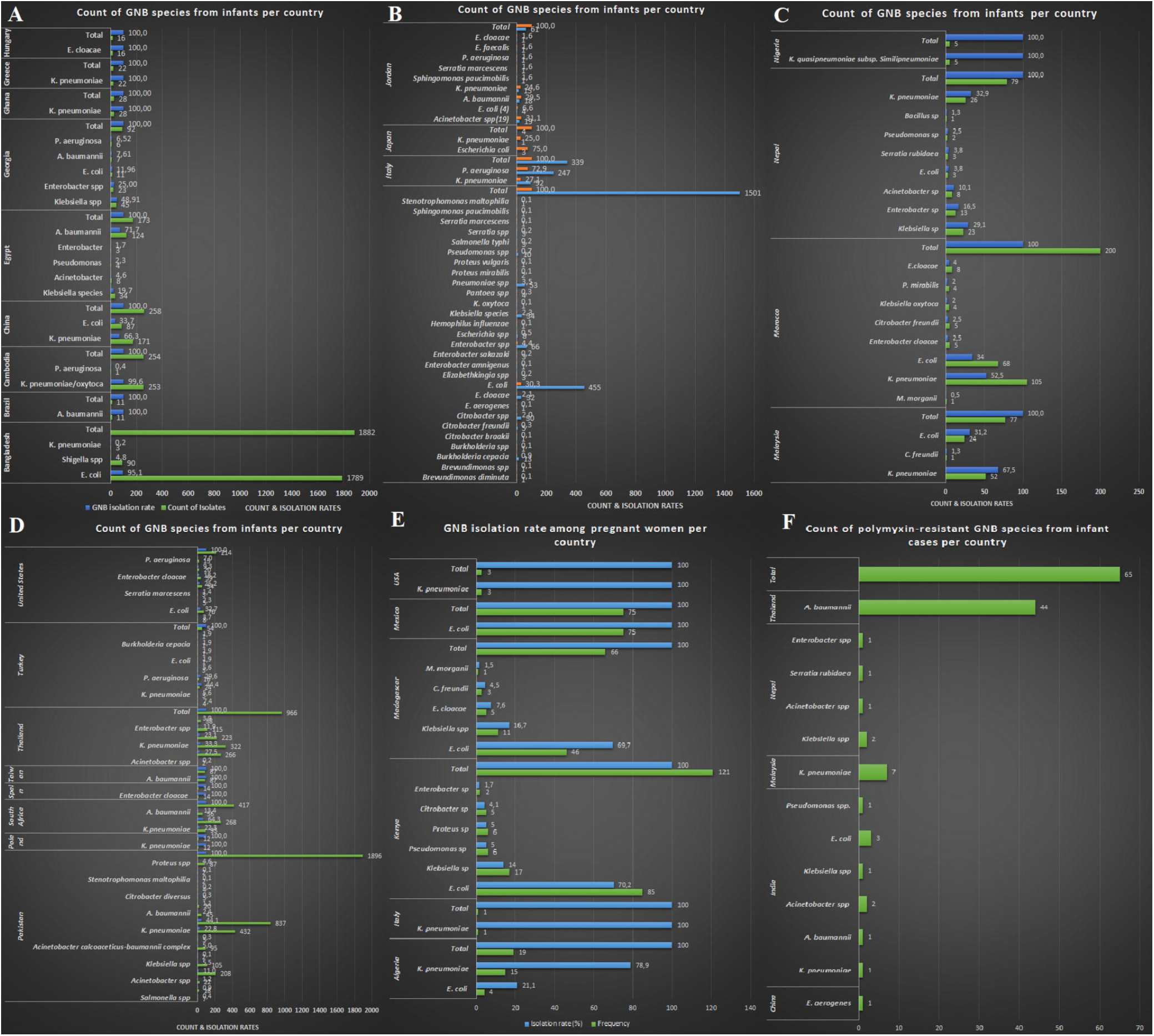
Count and isolation rates of Gram-negative bacterial species obtained from pregnant women and infants per country. **A-D** shows the count and isolation rates of carbapenem-resistant Gram-negative bacteria isolated from infants per country, **E** shows the count and isolation rates of CR-GNB in pregnant women whilst **F** shows the count of polymyxin-resistant GNB in infants.

The dominant clones of the various species were mainly localized within countries with few clones being observed across countries. For instance, only *K. pneumoniae* ST11 (in India, China, and multiple settings), *K. pneumoniae* ST15 (India, China, and Nepal), *K. pneumoniae* ST17 (Ghana and China) were observed across countries. Relatively few studies characterized the CR-GNB isolates to determine their clonality whilst others used PFGE or REP-PCR, limiting an effective international clonal analyses (Figure 2). Yet, the various clones identified within the countries were mostly involved in outbreaks in neonatal intensive care units (NICU) ^40–45^.

**Figure 2.**
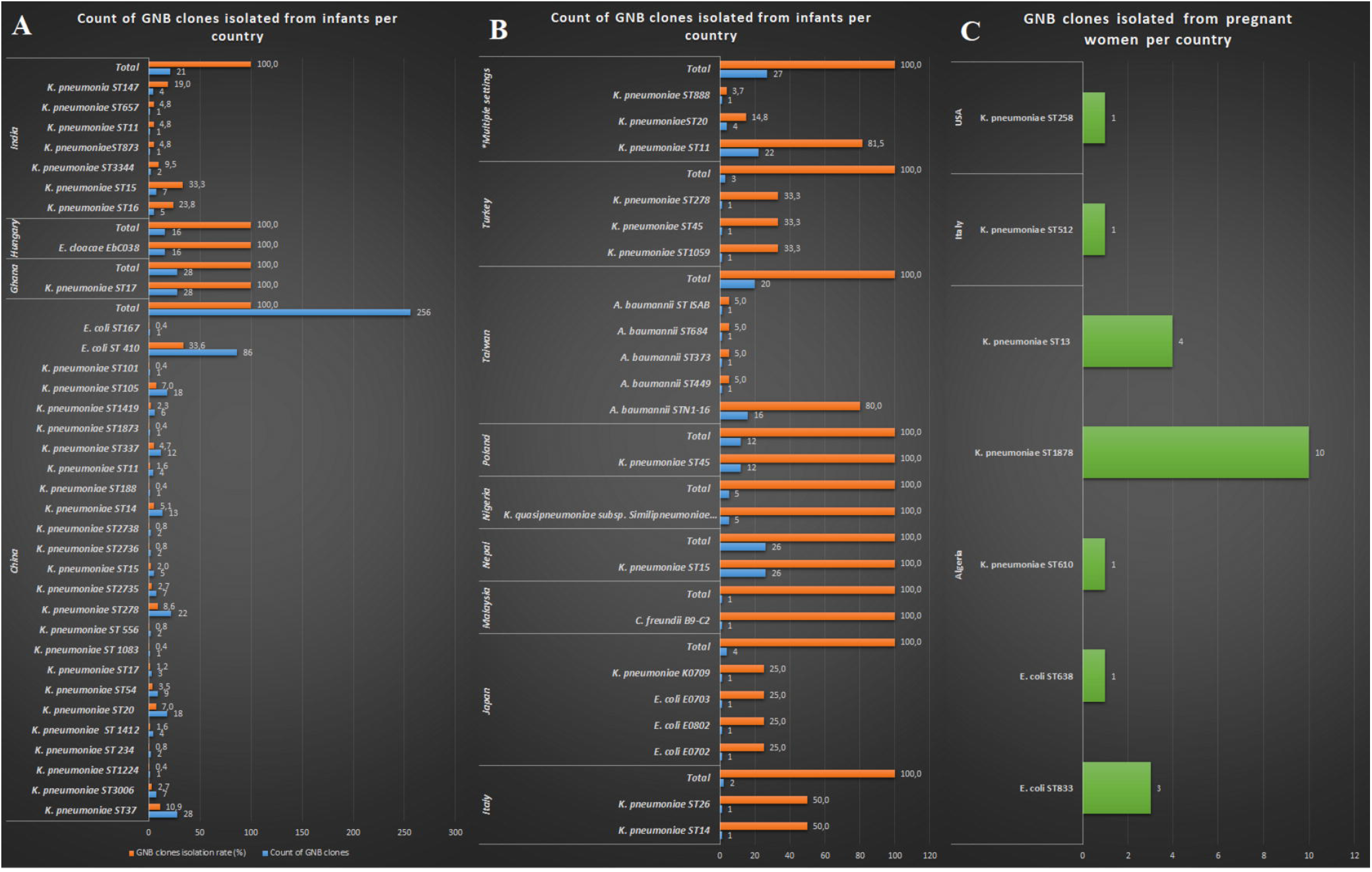
Count of Gram-negative bacteria clones infecting or colonizing infants and pregnant women per country. **A-B** shows count and prevalence of clones isolated from infants and **C** shows clones isolated from pregnant women.

Carbapenem resistance among infants in the various countries were substantial, particularly in India, Pakistan, China, Egypt, Taiwan, Thailand, and USA whilst polymyxin resistance rate in infants was high in Thailand, India, Malaysia, and Nepal. In pregnant women, Algeria, Madagascar, and Mexico recorded the highest number of CR-GNB colonizations (Figure 3).

**Figure 3.**
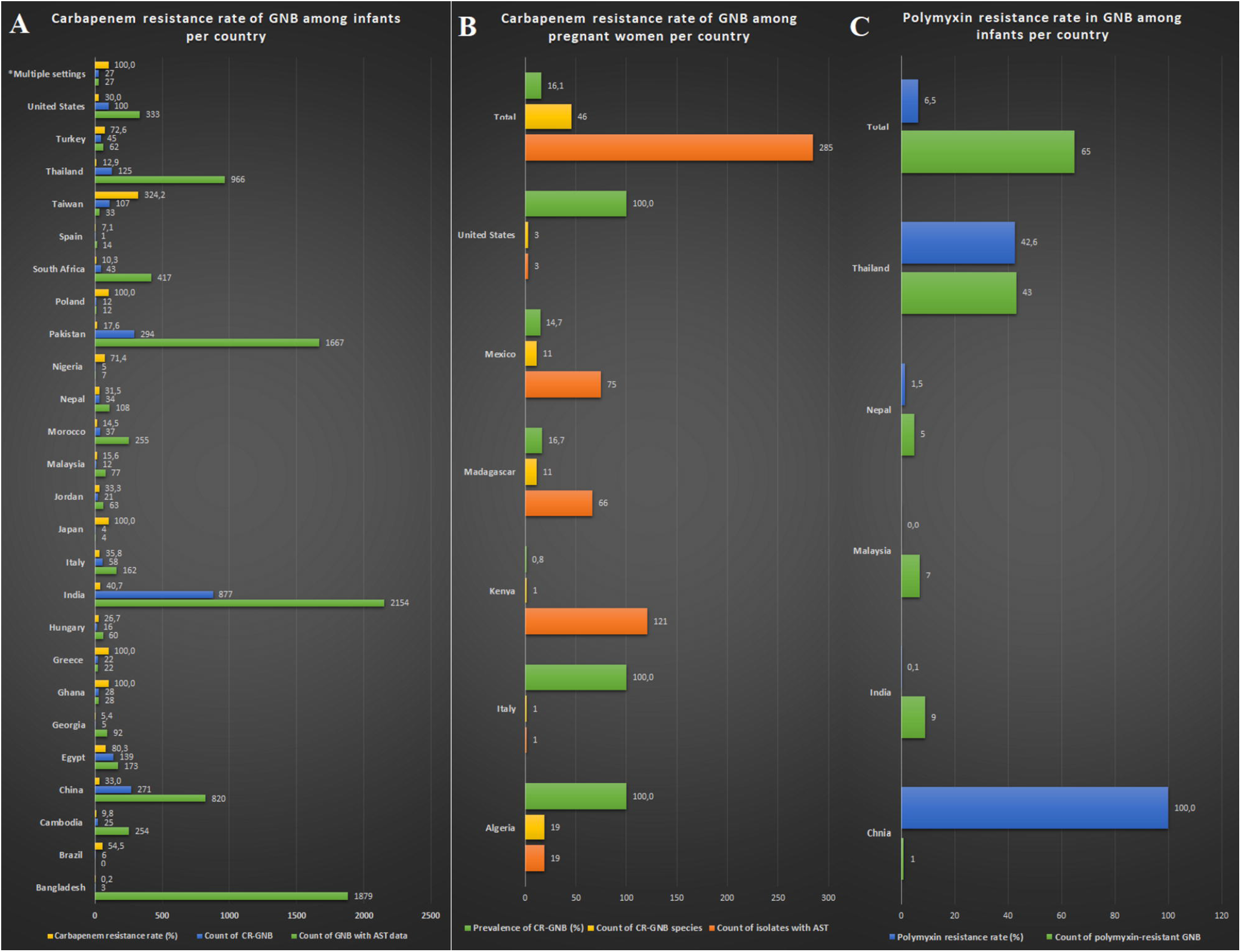
Carbapenem and polymyxin resistance rates in Gram-negative bacteria isolated from infants and pregnant women. **A** and **B** show carbapenem resistance rates from infants and pregnant women respectively, and **C** shows polymyxin resistance rates per country.

### Carbapenem & polymyxin resistance determinants

Carbapenem resistance among CR-GNB in both infants and pregnant women were mainly mediated by carbapenemases in all countries, with few reports of non-carbapenemases such as ompK35 and ompK36 porins loss (Italy), penicillin-binding protein mutations (PBP3) (Malaysia), OXA-1 β-lactamase (China), and soxS and marA efflux regulators (Malaysia), mediating carbapenem resistance. GES-5 (Poland), IMP (Japan and China), and VIM (Italy, Hungary, and Morocco) carbapenemases were mainly dominant in single or few countries or regions (such as the Mediterranean region) whilst NDM, KPC, and OXA-48-like variants were common in many countries among infants and pregnant women. Polymyxin resistance mechanisms were largely not described in the included studies, except in one study by Naha et al. (2020) in which overexpression of acrB, tolC, ramA, and soxS were found to mediate colistin resistance ^36^ and in another study by Zeng et al. (2016) in which *mcr-1* was found in an *Enterobacter* (now *Klebsiella*) *aerogenes* isolate (Figure 3).

### Risk factors and mortality rates

Risk factors associated with CR-GNB and PR-GNB infections and colonization differed slightly between countries, with factors such as previous and/or long hospitalization, previous and/or ongoing antibiotic therapy, invasive procedures such as caesarian section, intubation, and catheterization, preterm and low birth weight infants, and mechanical ventilation being common risk factors in almost all countries. Whereas PR-GNB-associated mortalities were not reported, CR-GNB-associated mortalities were above 10% in Brazil, China, Egypt, Hungary, and Thailand and noticeably lower than 10% in India, Nepal, Turkey, Pakistan, Italy, and Cambodia.

## Discussion

Carbapenem- and/or polymyxin-resistant GNB are common in neonates and pregnant women in several countries as either infections or colonizations, threatening the lives of the affected patients as CR-GNB and PR-GNB are associated with longer hospitalization and higher morbidities and mortalities ^29,46–48^. Worryingly, many of the studies involved in this review reported on clonal and polyclonal outbreaks and infections of CR-GNB or PR-GNB among neonates in NICUs or colonization of CR-GNB in pregnant women involving pathogenic GNB such as *A. baumannii, E. coli, Enterobacter sp*., *K. pneumoniae*, and *P. aeruginosa* (Tables S1-S3). In most of these CR-GNB, carbapenenemases, including NDM, OXA-48, KPC, VIM, IMP, and GES-5 were found on mobile plasmids within transposons, insertion sequences, and integrons.

Notably, carbapenem-resistant *A. baumannii, E. coli*, and *K. pneumoniae* harbouring mobile plasmid-borne carbapenemases cause outbreaks in neonates with sepsis, pneumonia, and other infections in NICUs in several countries worldwide, resulting in very high mortalities (Table S1) ^17,43,49–51^. For instance, carbapenem-resistant *A. baumannii* outbreaks were reported by Lee et al. (2018), Sultan and Seliem (2018), Indhar et al. (2017), Al-lawama et al. (2016), Thatrimontrichai et al. (2015), Kumar et al. (2014), and Tekin et al. (2013) in Taiwan, Egypt, Pakistan, Jordan, Greece, India, and Turkey respectively, killing several infants ^19,51–56^. *E. coli* neonatal infections and outbreaks were reported in Japan with an IMP-11 carbapenemase ^37^, in China with nine NDM-5-producing ST410 strains ^45^ and NDM-5-positive ST167^33^, in Turkey with an OXA-48-producing strain ^43^, and in India with NDM-1-positive clonal strains from the body of infants and the ward environment ^42^. Comparatively, outbreaks in NICUs involving *K. pneumoniae* were globally reported, involving NICUs infections, colonizations, mobile plasmid-borne *bla*_NDM_, *bla*_KPC_, *bla*_OXA-48_, *bla*_IMP,_ *bla*_VIM_ and *bla*_GES-5_ carbapenemases, and very high mortalities, making *K. pneumoniae* the most deadly and common neonatal MDR pathogen ^13,18,25,27,28,31,36,40,41,43,44,49,50,57–63^.

Similarly, PR-GNB infections were also common in the same species viz., *A. baumannii, E. coli, K. pneumoniae, Enterobacter sp*. and *Pseudomonas sp*., although reports on outbreaks were either associated with carbapenem resistance ^34,36^ or absent. Hence, outbreaks involving CR-GNB that were also polymyxin resistant could also be referred to as PR-GNB outbreaks, which makes such infections very difficult to manage ^1,2,4,9^. The largest collection of PR-GNB was recorded in Malaysia from preterm infants in a NICU in 2016, eight of which were carbapenem-resistant *K. pneumoniae* ^34^. A colistin-resistant *C. freundii* isolate was subsequently isolated from the stool of a preterm infant in the same hospital in Malaysia, suggesting the persistence of PR-GNB in that setting ^64^. Naha et al. (2020) observed that overexpression of acrB, tolC, ramA, and soxS conferred colistin resistance in a carbapenem-resistant *K. pneumoniae* ST147, which could have been the resistance mechanism in the *C. freundii* isolate from Malaysia ^36,64^. Although *mcr* genes, and not chromosomal mutations, are the most common mechanisms mediating polymyxin resistance globally ^1,2,4,9^, an *mcr-1* was only identified in one study (in *E. aerogenes*, now *K. aerogenes*) and interestingly, from the vaginal secretions of a pregnant woman ^39^.

Hence, vaginal swabbing could also be used to screen for CR/PR-GNB in pregnant women besides rectal swabbing as such important pathogens can be missed if they do not occur in the gut. Further, the presence of CR/PR-GNB in the vagina could explain the colonization of infants and wards with these MDR pathogens as they can pick up these bacteria during birth ^39,42^. If gut colonization of pregnant women with CR-GNB could potentially lead to colonization and/or infection of the neonate ^22,24,65^, then vaginal colonization could even lead to a more direct effect than gut colonization, making vaginal screening of pregnant women an important means of preventing CR/PR-GNB infections and mortalities in neonates. The means by which infants become colonized by CR/PR-GNB remains enigmatic, although direct inoculation from mothers is highly likely. Nevertheless, neonatal colonization with CR/PR-GNB is not only obtained from the mother as observed by other clinicians^20,23,26,42,61,66,67^.

In particular, neonates can become colonized and later infected with CR/PR-GNB during antibiotic therapy ^23,68^, long-stay in wards (NICU) with endemic CR/PR-GNB ^42,67^, handling by healthcare workers, breastfeeding by colonized mothers, intubation, during labour or caesarian section ^24,65,66^, mechanized ventilation, from the community ^23,61^ etc. (Tables S1-S3). To establish a direct relationship between mother and infant colonization, and with infant colonization and infection, then a clonal analyses of isolates would have to be undertaken. As yet, studies showing this clonal relationship between GNB isolates from mother and neonate, and between neonate colonization and infection, are wanting (Figure 2). Despite the larger number of studies reporting on pregnant women colonization, there are relatively few reports of pregnant women infection with CR/-PR-GNB, suggesting that colonization does not necessarily lead to infections in pregnant women or that their stronger and much developed immune system makes them less susceptible to CR/PR-GNB infections. Hence, CR/PR-GNB colonization in pregnant women is more of a risk to neonates than to the women themselves. Nevertheless, infection of pregnant women with CR/PR-GNB could complicate their pregnancies and put their lives at risk ^46^.

The clonality of the species infecting both pregnant women and infants were not the same, suggesting little dissemination between colonized mothers and neonates. Further, there were very few clones of *K. pneumoniae* that were seen in at most three countries, with almost all the reported clones of all the species being local and restricted to single hospitals and wards (Figure 2; Tables S1-S3). This demonstrates limited circulation of the same clone even within a community, ward, hospital or country.

The carbapenemases found in the CR-GNB isolates mirrored those found in the same countries and regions (Figure 4). For instance, IMP and VIM are common in Japan/China and Italy (Europe) respectively, OXA-48 is common in the mediterranean region, Middle-East and South Africa whilst KPC and NDM are globally distributed ^69^. This global epidemiology of carbapenemases is reflected in the epidemiology of carbapenemases in the infants and pregnant women (Figure 4), further corroborating the localized transmission of CR-GNB in neonates. In addition, the higher prevalence of NDM, KPC, and OXA-48-like carbapenemases over VIM and IMP ^69^, also reflects the global prevalence of these β-lactamases in neonates (Figure 4). The carbapenemases were mainly found on mobile plasmids (IncFII, IncX_3,_ IncN), integrons (*IntI*1), insertion sequences (*IS*Kpn7, *IS*Kpn6, *IS*Aba125, *IS*26) and transposons (Tn*4401b*), which facilitates their movement from chromosomes to plasmids, plasmids to plasmids, and between cells of the same or different species ^13,20,26,31,32,36,37,42,45,60,70^.

**Figure 4.**
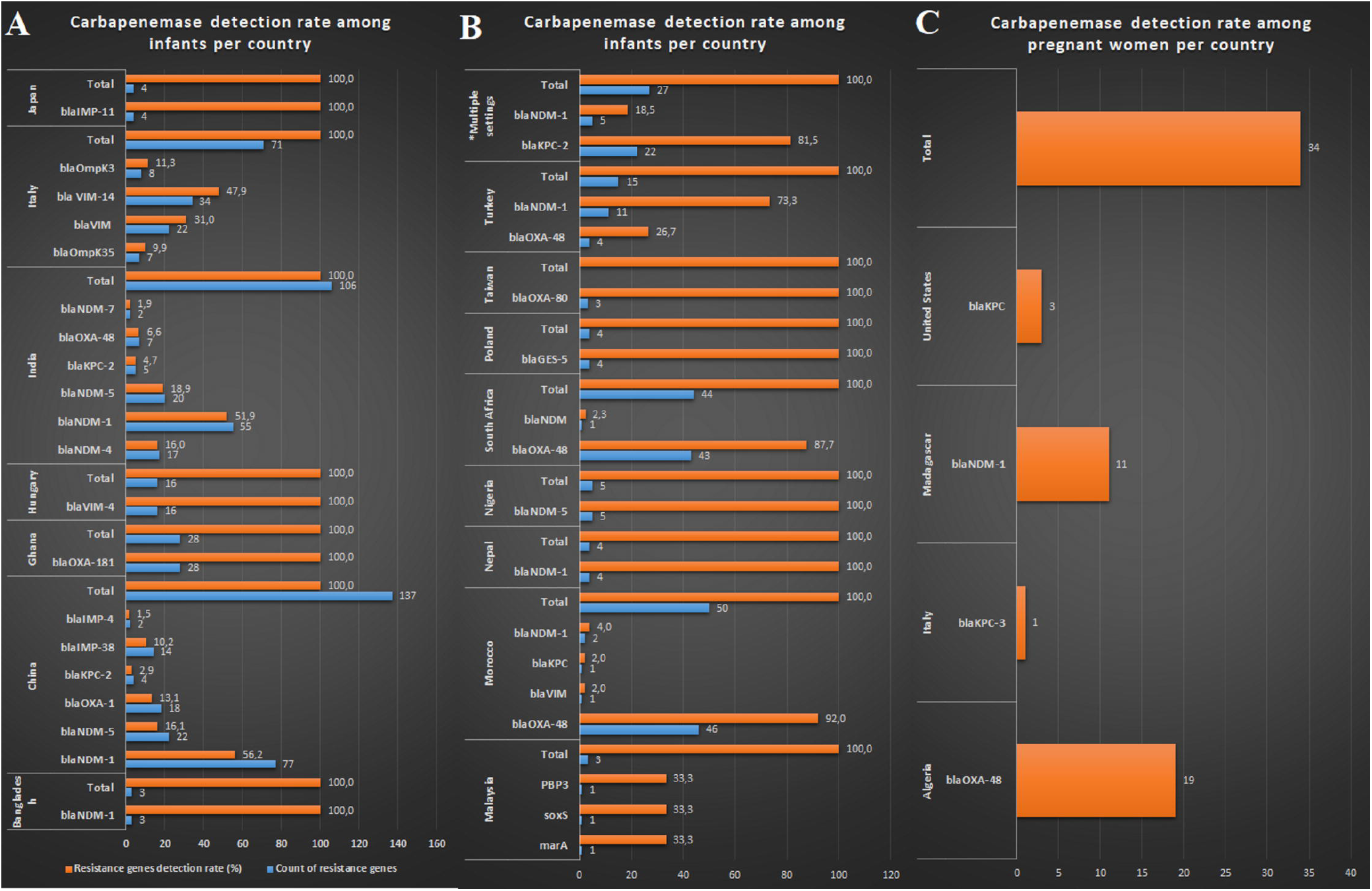
Carbapenemases detected in Gram-negative bacteria per country. **A** and **B** show carbapenemases detected in bacteria isolated from infants and **C** shows carbapenemases obtained from pregnant women.

Specifically, NDM-5 carbapenemases were found on IncX_3_ and IncFII plasmids in *K. quasipneumoniae* ST476 (in Nigeria) and *E. coli* ST410 (in China) respectively, which were causing outbreaks in NICUs. The immediate genetic environment of the NDM-5 on these plasmids were exactly the same or similar, suggesting that same plasmids and/or clones were mediating the spread of NDM-5 within and between species ^31,45^. The higher mobility of these carbapenemases may explain their faster dissemination between neonates in wards and hospitals during outbreaks and evinces the need to undertake molecular analyses of isolates to comprehend their epidemiology ^1,2,4,9,69^.

Not all CR-GNB isolates contained carbapenemases. Isolates without carbapenemases, but with ESBLs such as CTX-M, TEM, SHV, and OXA, coupled with the loss or downregulation of porins ompK35 and ompK36, had very high resistance to carbapenems ^27,44,59^. In Italy, such strains resulted in mortalities as high as 28.5% ^59^.

Prevalence of CR/PR-GNB and carbapenemases were higher in countries with higher GNB isolation rates (Figure 3-4), which is expected as most clinical investigations are undertaken in response to increasing infections or outbreaks. Further, susceptible GNB are hardly published except when they are isolated together with resistant GNB, making the literature skewed towards resistant GNB cases. Higher reports of carbapenemases were observed in China, India, Ghana, Italy, Hungary, Turkey, South Africa, Algeria and Madagascar (in pregnant women), which does not fully reflect the carbapenem resistance rates shown in Figure 3 as not all studies investigated the underlying carbapenem resistance mechanism. For instance, Cambodia, Egypt, Greece, Pakistan, Taiwan, Thailand, and USA had higher resistance rates, but very few or no report of carbapenemases. Whereas this may be due to financial limitations in low- and middle-income countries (LMICs), the same can not be true of wealthier countries such as Taiwan, Thailand, and the USA. Hence, there is the need for increasing awareness among clinicians and researchers on the need to undertake advanced genomics analyses of resistant GNB to facilitate epidemiological analyses and interventions.

On the other hand, countries with higher prevalence of carbapenemases and carbepnem resistance rates should immediately institute effective infection control and prevention practices as sufficient evidence indicates that this can both prevent and contain CR/PR-GNB outbreaks ^41,63^. Particularly, the nosocomial colonization of neonates with clonally related carbapenemase-producing *P. aeruginosa* in a NICU and the subsequent isolation of a VIM-14 carbapenemase in the same pathogen in a NICU, both in Italy ^71,72^, depict the endemic presence of this carbapenemase-producing pathogen within those periods in the affected regions. This signals the urgency required in instituting effective interventions to clear these MDR pathogens and save further lives as outbreaks in NICUs are associated with very high mortalities ^29,50,54^.

Common risk factors for getting CR/PR-GNB infections across all countries have been stated above: they include include colonization of the infant or mother with CR/PR-GNB, long-term hospitalization in NICUs, undergoing invasive procedures and previous, ongoing or longer antibiotic therapy, mechanical ventilation, preterm state, and low birth weight (Fig. 5) ^22,55,57^. Interestingly, neonatal colonization with CR/PR-GNB during NICU stay can later result in community and nosocomial infections as described by Vergadi et al. (2017) ^61^. Sadly, infant mortalities from CR/PR-GNB remains substantially high in many countries, including China and Brazil (Fig. 6), albeit treatment with carbapenems (at higher doses), colistin, and tigecycline have been shown to be very efficient in clearing the infection and saving lives ^7,10,28,45,56,73^. Instructively, Kumar et al. (2014) observed that the absence of effective treatment led to higher all-cause mortalities in neonates ^54^.

**Figure 5.**
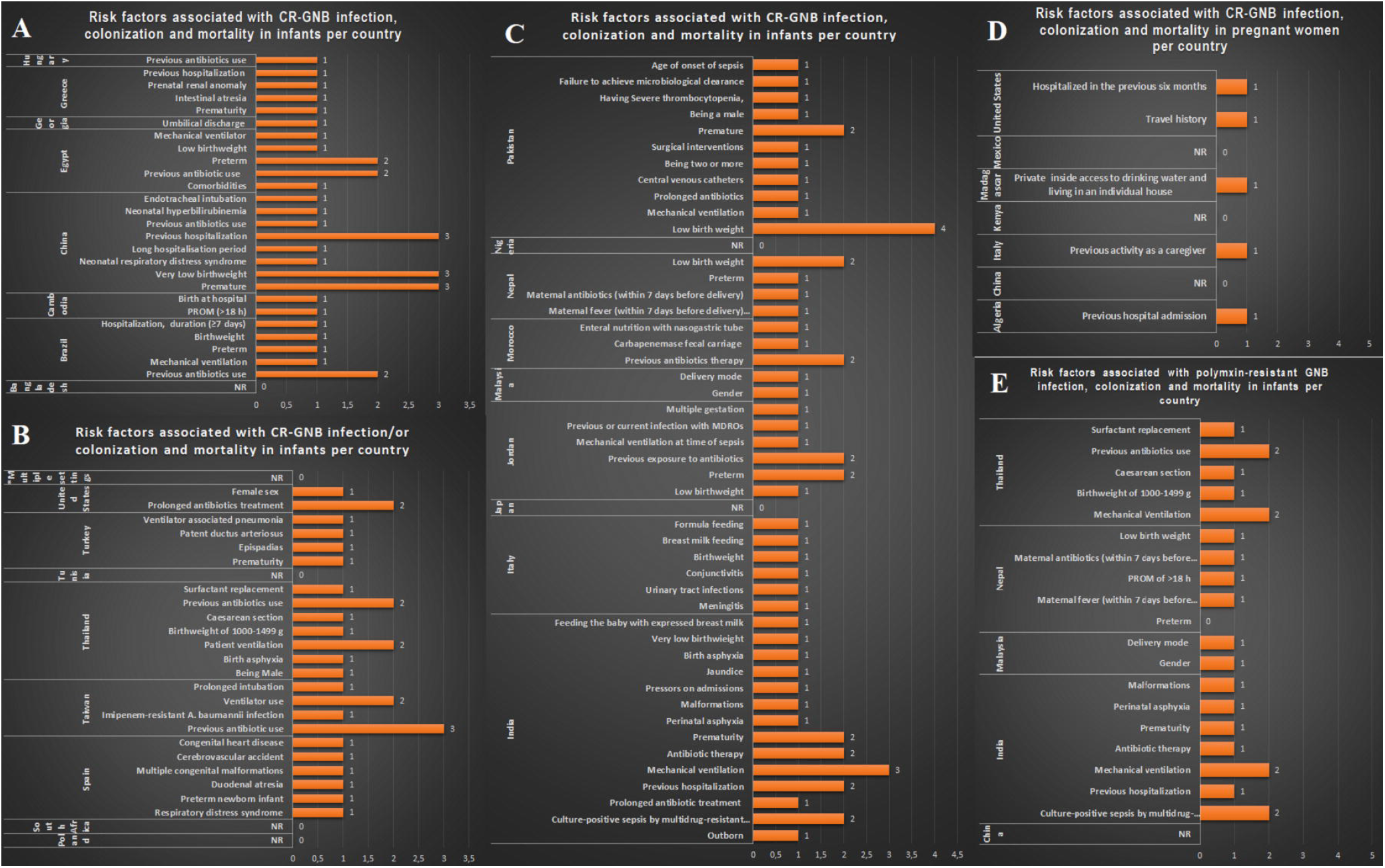
Risk factors associated with carbapenem and polymyxin-resistant Gram-negative bacteria in neonates and pregnant women. **A-C** depict risks associated with carbapenem resistance in neonates, **D** depict risks associated with carbapenem resistance in pregnant women, and **E** depicts risks associated with polymyxin/colistin resistance in neonates and pregnant women.

**Figure 6.**
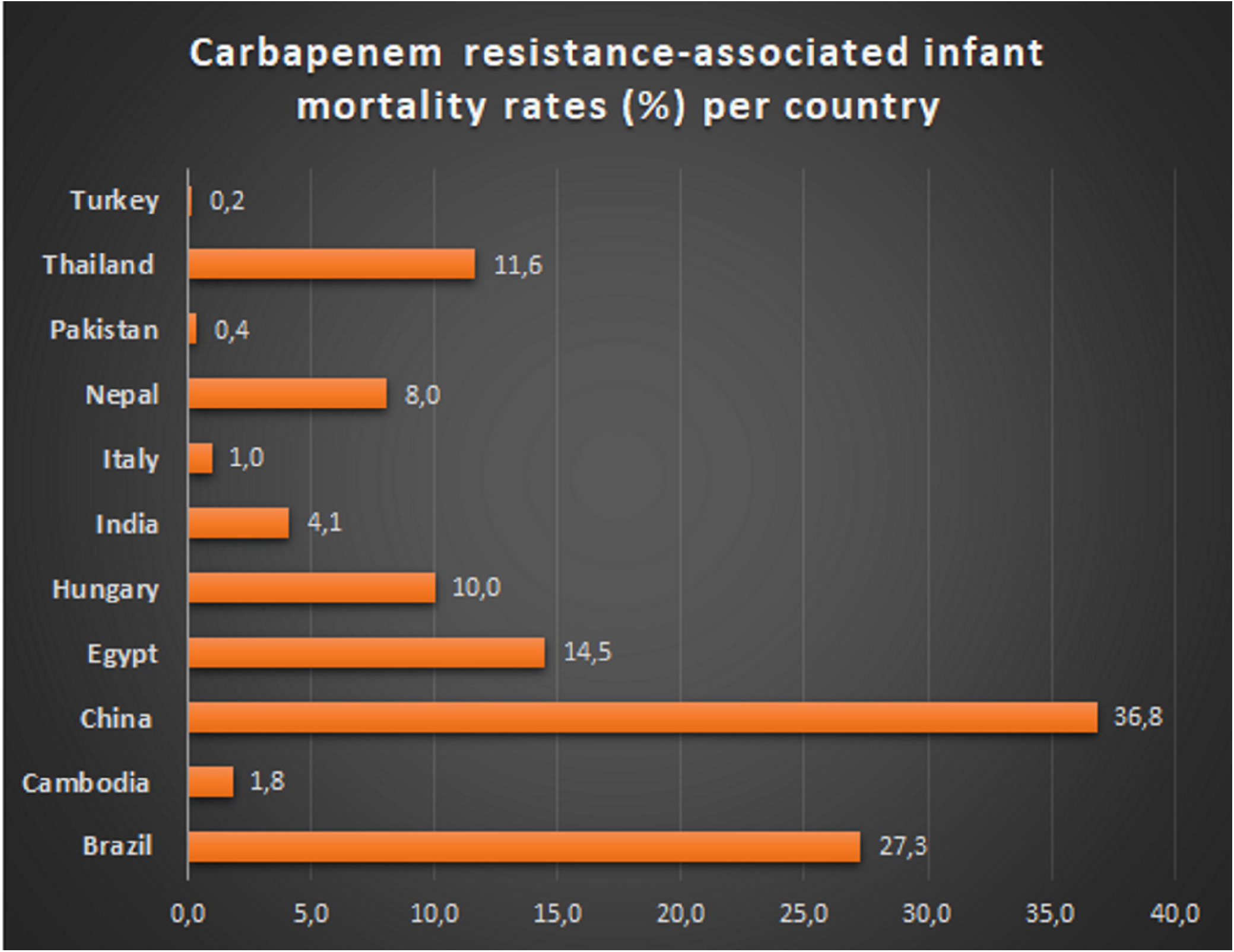
Mortality rates associated with carbapenem resistance in infants per country.

### Conclusion and future perspectives

Evidently, carbapenem and polymyxin resistance is common in GNB isolated from infants and pregnant women, with high resistance rates and diverse carbapenemases globally, particularly in India, China, Pakistan, Thailand, Algeria, South Africa, Turkey, Ghana, Hungary, and Italy. In almost all these countries, there were local clonal outbreaks involving *A. baumanii, E. coli, Enterobacter sp*., *K. pneumoniae*, and *P. aeruginosa* clones hosting mobile plasmids, integrons, insertion sequences, and transposons on which were found *bla*_NDM_, *bla*_KPC_, *bla*_OXA-48_, *bla*_IMP,_ *bla*_VIM_ and *bla*_GES-5_ carbapenemases. Whereas colonization in pregnant women is a risk factor for neonatal infections and outbreaks, infections in pregnant women can complicate their pregnancies. Infection with CR/PR-GNB has been associated with very high mortality rates among infants, necessitating urgent infection control interventions to prevent further outbreaks and safeguard lives. The commonality of risk factors for getting CR/PR-GNB infections in all countries indicate that adopting a standard and strict infection control measures in all healthcare centres globally can contain these pathogens from causing further infections. Several outbreaks have been contained with strict infection control interventions.

Fortunately, treatment of CR/PR-GNB infections is possible with polymyxins, higher doses of carbapenems, tigecycline, amikacin, and avibactam/sulbactam-cephalosporin/carbapenem combinations. Such treatment protocols and medicines must be distributed to all paediatricians and enforced to reduce infant mortalities. Nevertheless caution with antibiotic therapy (stewardship) must be also advised as it is a risk factor for acquired carbapenem resistance colonization or infection.

Going forward, periodic genomic (molecular) surveillance/screening of hospitals (wards) and patients must be instituted to pre-emptively identify patients with CR/PR-GNB infections or colonization. Particularly, vaginal and rectal screening of pregnant women can help in surveillance exercises. Further genomic analyses to identify the clones, plasmids, and resistance mechanisms shall inform the appropriate intevention to adopt to break the chain of transmission.

## Data Availability

All data are included as supplementary material

## Acknowledgments

None

## Transparency declaration

None

---

**Figure S1. Literature search strategy, inclusion and exclusion criteria employed to arrive at the articles used in this study**.

**Supplemental Table 1**. Data on the epidemiology, risk factors, clinical outcomes, resistance mechanisms, resistance rates, and prevalence of carbapenem-resistant Gram-negative bacteria infecting or colonizing infants.

**Supplemental Table 2**. Data on the epidemiology, risk factors, clinical outcomes, resistance mechanisms, resistance rates, and prevalence of carbapenem-resistant Gram-negative bacteria infecting or colonizing pregnant women.

**Supplemental Table 3**. Data on the epidemiology, risk factors, clinical outcomes, resistance mechanisms, resistance rates, and prevalence of polymyxin-resistant Gram-negative bacteria infecting or colonizing infants and pregnant women.

**Figure S1:**
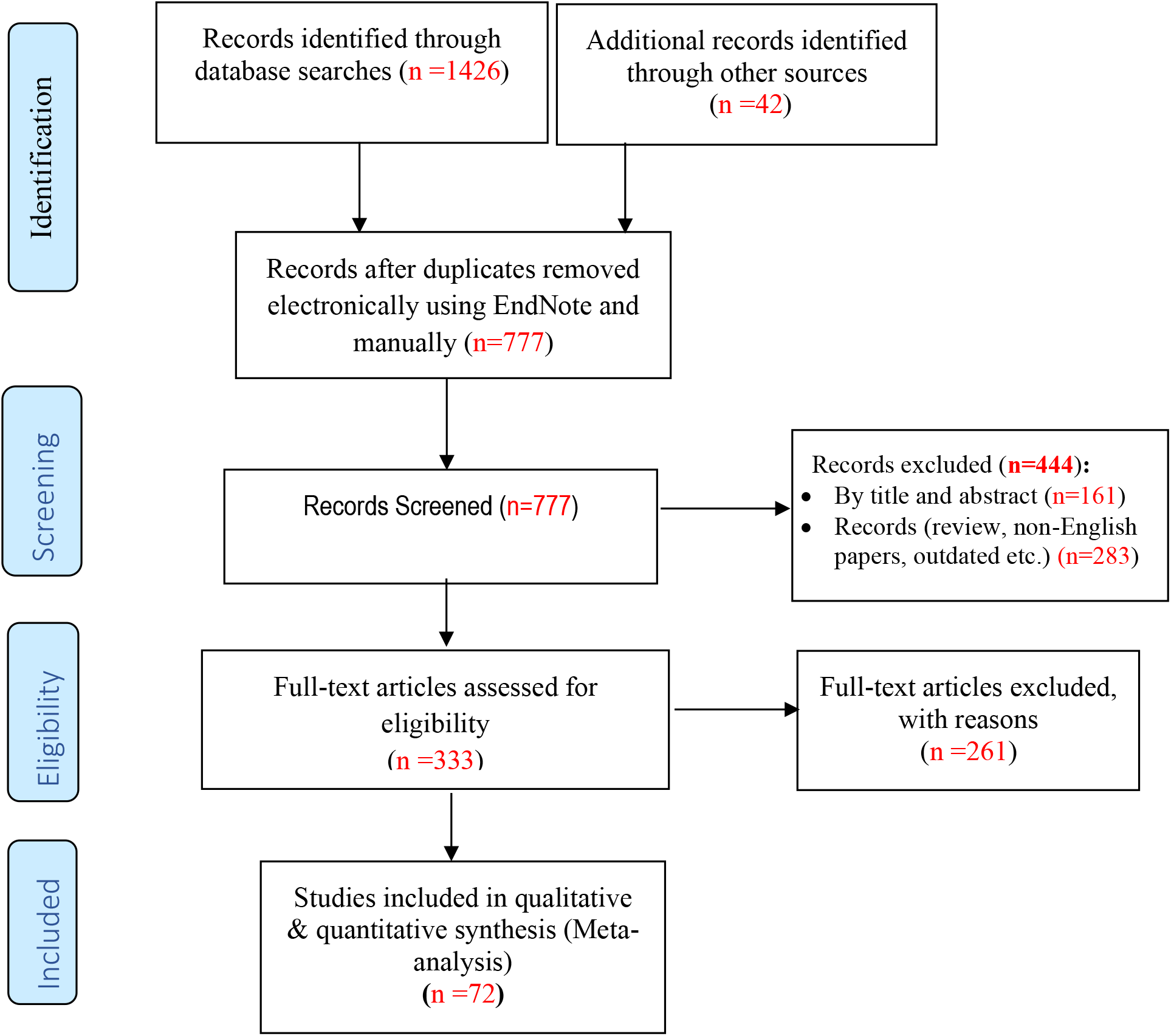
Flow diagram showing the results of the literature and database search strategy, inclusion, and clusion criteria, with reasons.

**Figure S2.**
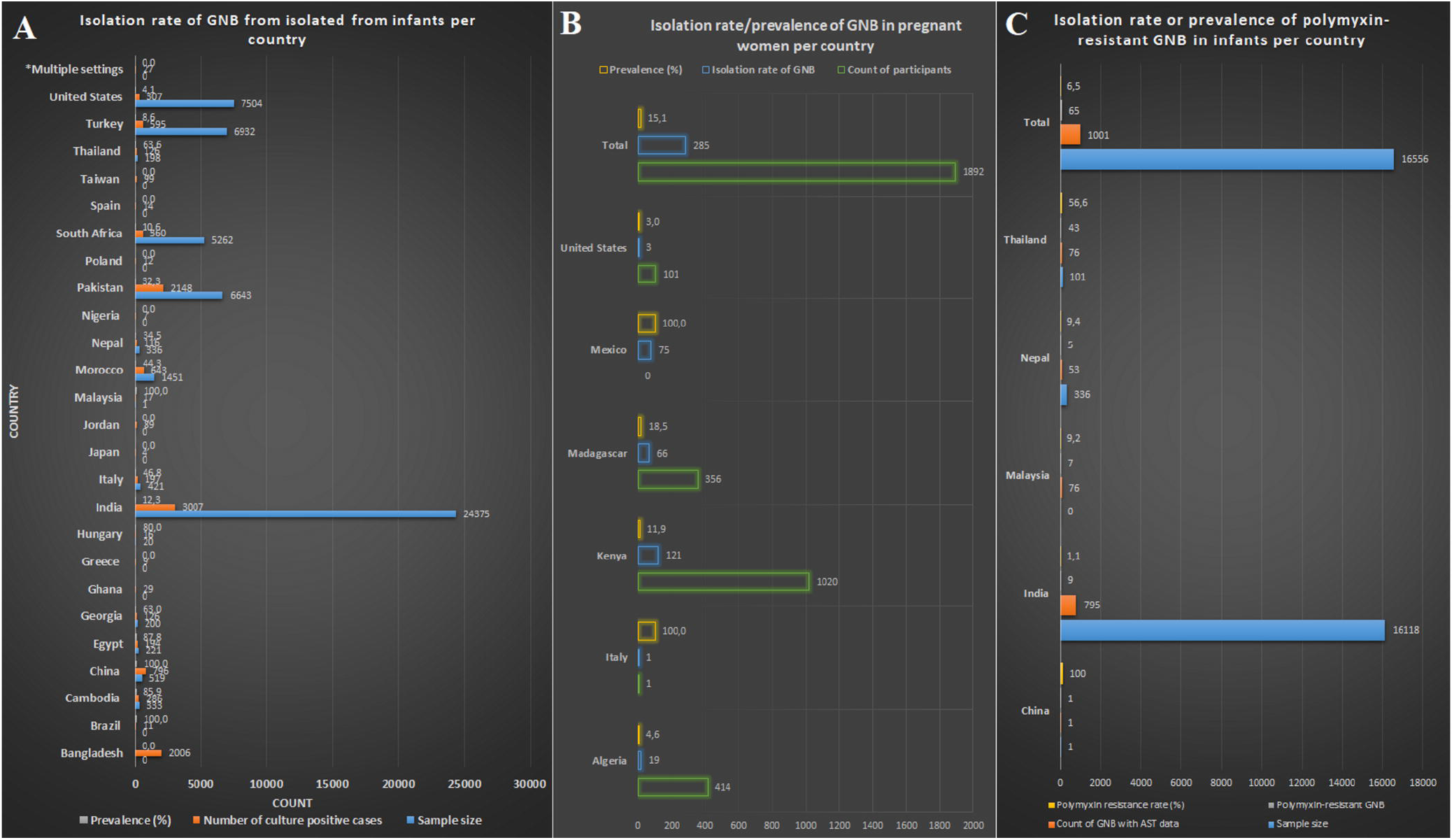
Isolation rate of Gram-negative bacterial species obtained from infants and pregnant women per country. **A** shows the isolation rate of Gram-negative bacteria obtained from infants per country, **B** shows the isolation rate of Gram-negative bacteria obtained from pregnant women per country, and **C** shows the isolation rate of polymyxin-resistant Gram-negative bacteria obtained from infants per country,

